# The Nova Protocol: A Comprehensive Multimodal Longitudinal Study among Civilian Survivors of a Mass Trauma Event

**DOI:** 10.64898/2026.08.09.26359968

**Authors:** Roee Admon, Ophir Netzer, Noa Magal, Lisa Simon, Amir Harduf, Michal Oren, Omri Radai, Stephanie Keren Cohen, Meital Bobek, Mary Grankin, Ron Menshes, Yonatan Stern, Noga Mandelblit, Avraham Shmueli, Eden Eldar, Daniel Sand, Tzuk Polinsky, Raz Gross, Roy Salomon

## Abstract

**Background:** The October 7, 2023 attack in southern Israel was one of the deadliest terror attacks in modern history, with 1,182 fatalities, more than 4,000 wounded individuals, and 251 hostages. The Nova music festival, an all-night outdoor rave near the Gaza border, suffered the highest number of civilian casualties, with more than 370 festival attendees killed. Survivors were exposed to prolonged life-threatening trauma with similar characteristics and within a narrow time window. Many survivors also reported being under the acute influence of psychoactive substances during the attack and the following hours. This tragic combination of civilian mass trauma and naturalistic pharmacological exposure created a rare opportunity to study trauma processing prospectively.

**Objective:** This paper describes the rationale, design, and methodology of the Nova Protocol, a multimodal longitudinal observational study of survivors of the October 7, 2023 Nova festival attack and a sociocultural comparison group.

**Methods:** The protocol spans from the first weeks to approximately 24 months post-trauma and includes three major assessment time points. It integrates repeated online clinical assessments, prolonged wearable-sensor monitoring, ecological assessments, saliva-based endocrine and inflammatory markers, structural and functional MRI, cardiac interoception paradigms, online and in-scanner reinforcement-learning tasks, and semi-structured qualitative interviews. Primary outcomes are PTSD symptom severity (PCL-5) and general psychological distress (K6), supplemented by a rich battery of secondary measures.

**Conclusion:** The Nova Protocol provides an unusually rich longitudinal framework for characterizing psychological, behavioral, physiological, inflammatory, neural, interoceptive, and subjective mechanisms that shape clinical trajectories after civilian mass trauma. Because psychoactive substance exposure was naturalistic and self-selected, findings will be interpreted as mechanistic and prognostic associations rather than causal effects. The protocol is expected to inform early risk detection and scalable post-disaster monitoring and intervention strategies, as well as unique insights into how psychoactive substances impact trauma processing.

**HIGHLIGHTS:**

- We describe a multimodal longitudinal protocol tracking long-term responses to severe trauma among civilian survivors of the October 7, 2023 Nova festival attack.
- The protocol leverages a rare naturalistic cohort of participants who were exposed to the same mass trauma event within a narrow time window, with many survivors also reporting being under the influence of psychoactive substances during the attack.
- Survivors were longitudinally assessed starting from the first weeks post-trauma to approximately 24 months later.
- Assessments integrate repeated clinical questionnaires, wearable sensors, ecological assessments, saliva-based endocrine and inflammatory markers, structural and functional MRI, cardiac interoception paradigms, behavioral reinforcement-learning tasks, and semi-structured qualitative interviews.
- The protocol enables multi-level investigation of trauma responses in a naturalistic mass trauma context, with implications for early risk detection, post-disaster mental health care, and the study of peritraumatic psychoactive pharmacology.

## 1. Background

Post-traumatic stress disorder (PTSD) is a prevalent and disabling consequence of exposure to severe trauma such as interpersonal violence, terrorism, and mass-casualty events (Charlson et al., 2019; Paz Garcia-Vera et al., 2016; Rigutto et al., 2021; Shalev et al., 2017). Trauma responses are heterogeneous, and longitudinal studies commonly identify fluctuations in symptom severity over time that may be expressed as resilient, recovery, chronic, and delayed/worsening symptom trajectories (Galatzer-Levy et al., 2018; Lowe et al., 2021; Simon et al., 2026). The peritraumatic period, defined as the interval during and immediately following trauma exposure, is increasingly regarded as a critical window in which biological, psychological, and social processes interact to shape long-term recovery or chronicity (Brewin et al., 2000; Bryant, 2011; Ozer et al., 2003). Identifying early markers of chronic PTSD risk has substantial clinical value because it may support the allocation of monitoring and treatment resources to survivors most in need.

A large body of literature has identified the neural, biological, physiological, cognitive, and psychosocial correlates of PTSD (Brewin et al., 2000; Ozer et al., 2003; Pitman et al., 2012; Quinones et al., 2020; Rab & Admon, 2021; Ressler et al., 2022; Scott et al., 2015; Shalev et al., 2017). However, much of this evidence is based on cross-sectional studies of chronic PTSD patients or on samples that combine different trauma types and variable intervals from trauma exposure to assessment. Such designs make it difficult to characterize peritraumatic mechanisms, temporal ordering, and evolving symptom trajectories. One exception is the extensive longitudinal study of the September 11, 2001 World Trade Center (WTC) attacks. By enrolling over 71,000 rescue/recovery workers and community members, the WTC Health Registry demonstrated that PTSD trajectories are heterogeneous and long-lasting, with a 20-year follow-up finding persistent or worsening of PTSD symptoms in approximately 10% of responders (Mann et al., 2025). Notably, even these large-scale efforts often relied primarily on self-report assessments and began data collection well after the acute post-trauma period (Ko et al., 2022). Prospective studies initiated in the immediate aftermath of trauma exposure are therefore essential for distinguishing early predictors, correlates, and consequences of PTSD development (Admon et al., 2013; Ben-Zion et al., 2019).

The October 7, 2023 attack in southern Israel resulted in 1,182 fatalities, more than 4,000 wounded individuals, and 251 individuals taken hostage, making it one of the deadliest terror attacks in modern history (Levi-Belz et al., 2024). The Nova music festival, an overnight trance rave held near the Gaza border alongside several smaller raves, sustained the highest number of civilian casualties during the attack, with more than 370 festival attendees killed. The remaining survivors endured prolonged exposure to acute, severe, life-threatening traumatic events, often over many hours. Furthermore, the majority of survivors reported being under the influence of psychoactive substances during the attack, and most of these participants reported substance consumption within the three hours preceding attack onset (Netzer et al., 2025). This tragic confluence of civilian mass trauma and naturalistic pharmacological exposure created a rare naturalistic research opportunity.

The Nova Protocol was designed to leverage this unique naturalistic cohort in order to address existing knowledge gaps in the literature. It does so through a multimodal, longitudinal study initiated with 1,268 eligible Nova survivors that began in the early aftermath of the event and extended up to approximately two years post-trauma, with modality-specific sampling across time points. By integrating repeated self-reports, functional neuroimaging, daily-life wearable physiology, ecological assessments, saliva-based endocrine and inflammatory markers, cardiac interoception paradigms, reinforcement-learning tasks, and qualitative interviews, the protocol enables both population-level characterization of response trajectories starting from immediately after exposure, as well as mechanistic investigation of individual differences in trauma processing. A further unique feature of this cohort is the high prevalence of naturalistic peritraumatic psychoactive-substance exposure. Given the known effects of psychoactive substances, including MDMA and classic psychedelics, on fear processing, emotional regulation, sense of self and reality, social cognition, autonomic function, and inflammatory pathways (Drori et al., 2025; Flanagan & Nichols, 2018; Harduf et al., 2023; Kamilar-Britt & Bedi, 2015; Mitchell et al., 2023; Mithoefer et al., 2018; Nichols, 2022), this protocol offers an unusual opportunity to study peritraumatic pharmacology in an ethically non-replicable real-world context.

## 2. Methods

### 2.1 Study design

The Nova Protocol is a longitudinal observational study with three primary time points (**TP1**, **TP2**, **TP3**), spanning from the first weeks after the October 7, 2023 attack to approximately 24 months later. It includes two groups: a trauma-exposed group comprising direct survivors of the Nova festival attack (n = 1,268) and a comparison group comprising members of the Israeli trance music community who were not directly exposed to the October 7 attacks (n = 508). The sequential study stages integrate remote online assessments, prolonged daily-life monitoring, and in-person laboratory sessions. The study is observational: psychoactive substance exposure was naturalistic and self-selected, not assigned or manipulated by the research team. A schematic representation of the study timeline and assessment components is presented in **Figure 1**.

**Figure 1.**
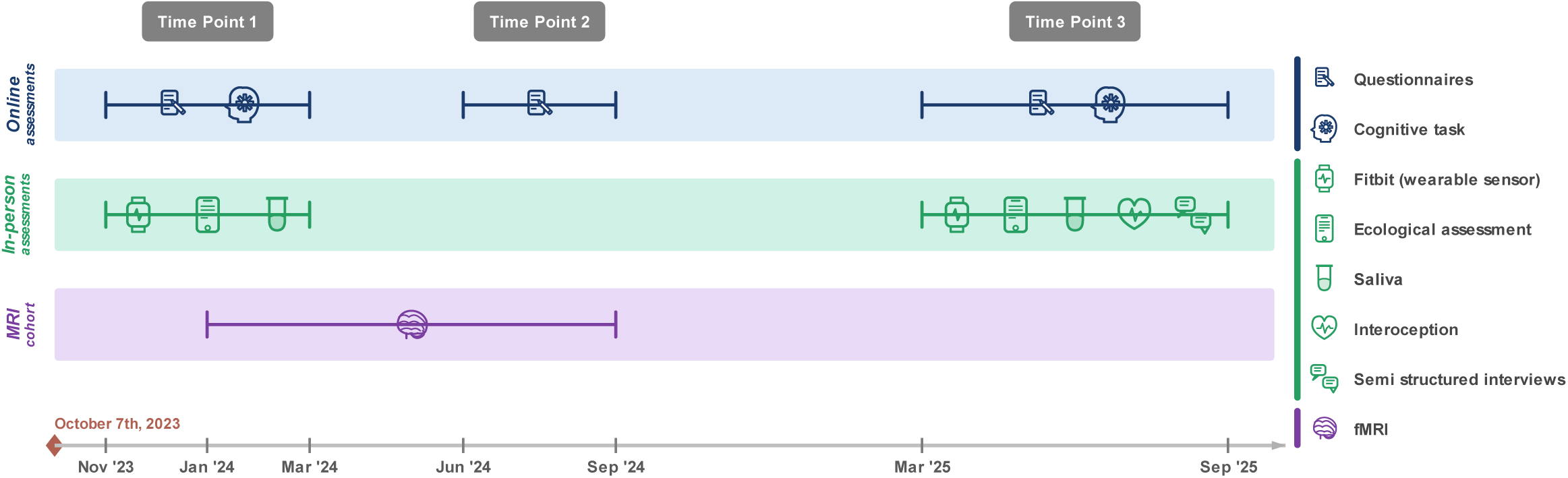
Schematic timeline of the Nova Protocol study design. Three assessment time points (Time Point 1, Time Point 2, Time Point 3) are shown relative to the October 7, 2023 traumatic event. Comparison participants were collected through Jan 2026. Modalities assessed at each time point include online questionnaires, Fitbit wearable sensors, ecological assessments, saliva-based endocrine and inflammatory markers, fMRI, cardiac interoception, reinforcement-learning tasks, and semi-structured interviews.

### 2.2 Participants

Nova survivors were individuals who were directly present at the Nova festival area during the October 7, 2023 attack. Survivors were recruited through SafeHeart, a non-profit organization established to support survivors of the Nova attack, peer referrals, and survivor-focused social-media support groups. The comparison group comprised individuals who attended other parties that were not attacked on October 7, 2023, or who regularly attend Israeli trance music festivals but were not present at the targeted events. This criterion referred to direct presence at, or direct life-threatening exposure to, the targeted October 7 events; indirect, vicarious, or subsequent war-related exposures were not exclusionary and were assessed separately. This design enables comparison between individuals directly exposed to trauma and non-exposed individuals from a shared sociocultural background, while accounting for putative pre-existing differences in festival attendance, lifestyle, and substance-use patterns.

### 2.3 Ethics approval, informed consent, and participant safety

The study was approved by the University of Haifa Institutional Ethics Committee (approval no. 374/23). The fMRI component was also approved by the Rambam Health Care Campus Institutional Review Board (RMB0553-23). The interoception sub-study as part of TP3 was preregistered on AsPredicted (registration #282,676). All participants provided informed consent before participation at each assessment time point. Given the sensitivity of the population, research assistants were instructed to emphasize voluntariness, avoid pressuring participants, and prioritize participant well-being. Participants who expressed distress or were suspected of experiencing distress were referred to SafeHeart or other relevant support organizations.

### 2.4 Eligibility criteria

Eligibility for both groups required age 18 years or older, Hebrew-language proficiency, and capacity to provide informed consent. For the fMRI subsample, additional exclusion criteria included current or prior diagnosis of neurological or psychiatric disorders, including PTSD prior to October 7, and MRI contraindications. Participants in the comparison group were required to confirm no direct exposure to the October 7 attacks.

### 2.5 Recruitment and timeline

Participant outreach for the Nova survivor group began on November 2, 2023, approximately 26 days post-event. Participants were contacted through dedicated smartphone applications, emails, text messages, and telephone calls. Remote assessments were administered via the Qualtrics online survey platform and, when needed, trained research staff assisted survivors in completing the online questionnaires by telephone. In-person sessions were conducted at the University of Haifa and affiliated sites, including designated community spaces and home visits for participants unable or unwilling to attend laboratory visits. For survivors, major assessment windows were TP1 (November 2023-March 2024), TP2 (June-September 2024), and TP3 (March-September 2025). For the comparison group, TP1 data were collected between November 2023 and April 2024, TP2 between June and September 2024, and TP3 between August 2025 and January 2026.

### 2.6 Study stages

Study stages are summarized in **Table 1** and described in detail below.

**Table 1.**
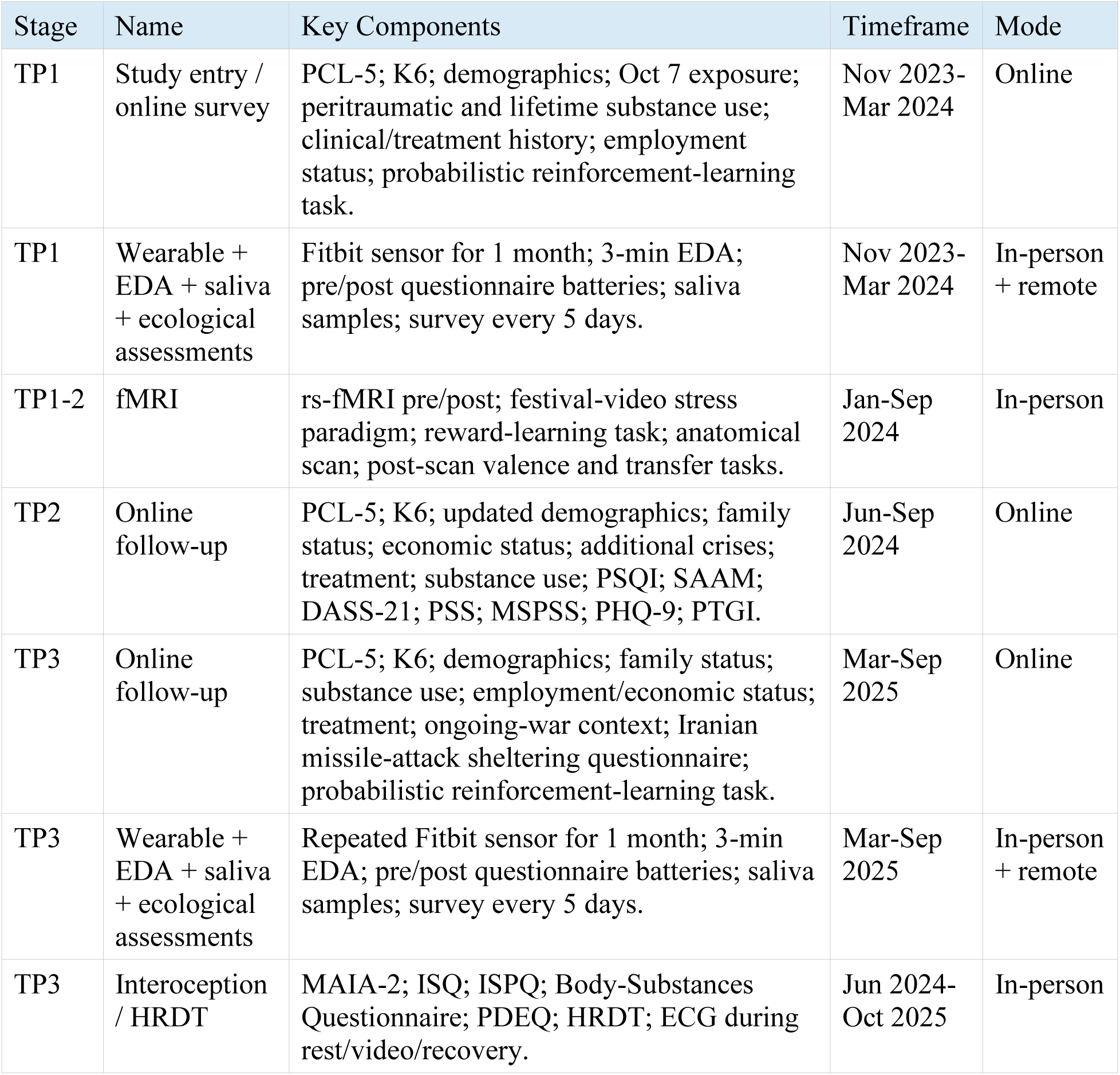
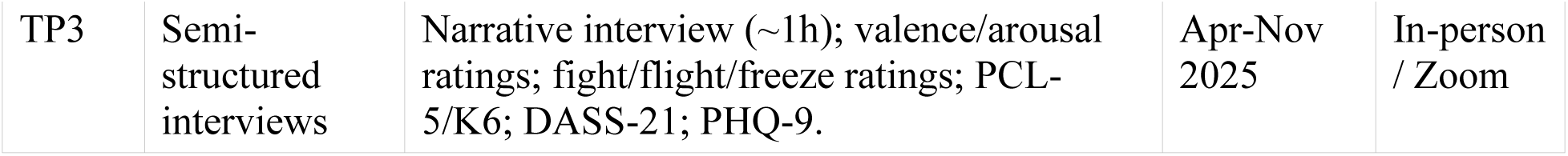
Study stages including all Time Points (TP).

| Stage | Name | Key Components | Timeframe | Mode |
| --- | --- | --- | --- | --- |
| TP1 | Study entry / online survey | PCL-5; K6; demographics; Oct 7 exposure; peritraumatic and lifetime substance use; clinical/treatment history; employment status; probabilistic reinforcement-learning task. | Nov 2023-Mar 2024 | Online |
| TP1 | Wearable + EDA + saliva + ecological assessments | Fitbit sensor for 1 month; 3-min EDA; pre/post questionnaire batteries; saliva samples; survey every 5 days. | Nov 2023-Mar 2024 | In-person + remote |
| TP1-2 | fMRI | rs-fMRI pre/post; festival-video stress paradigm; reward-learning task; anatomical scan; post-scan valence and transfer tasks. | Jan-Sep 2024 | In-person |
| TP2 | Online follow-up | PCL-5; K6; updated demographics; family status; economic status; additional crises; treatment; substance use; PSQI; SAAM; DASS-21; PSS; MSPSS; PHQ-9; PTGI. | Jun-Sep 2024 | Online |
| TP3 | Online follow-up | PCL-5; K6; demographics; family status; substance use; employment/economic status; treatment; ongoing-war context; Iranian missile-attack sheltering questionnaire; probabilistic reinforcement-learning task. | Mar-Sep 2025 | Online |
| TP3 | Wearable + EDA + saliva + ecological assessments | Repeated Fitbit sensor for 1 month; 3-min EDA; pre/post questionnaire batteries; saliva samples; survey every 5 days. | Mar-Sep 2025 | In-person + remote |
| TP3 | Interoception / HRDT | MAIA-2; ISQ; ISPQ; Body-Substances Questionnaire; PDEQ; HRDT; ECG during rest/video/recovery. | Jun 2024-Oct 2025 | In-person |
| TP3 | Semi-structured interviews | Narrative interview (~1h); valence/arousal ratings; fight/flight/freeze ratings; PCL-5/K6; DASS-21; PHQ-9. | Apr-Nov 2025 | In-person / Zoom |

#### TP1

- <u>Study entry:</u> Study entry comprised an online questionnaire battery administered between November 2023 and March 2024. The assessment captured demographics, pre-event clinical history, prior trauma exposure, substance-use history before October 7, detailed peritraumatic substance use during the attack, trauma-exposure severity, current PTSD symptom severity, psychological distress, mental-health treatment history, measures of social and emotional functioning and employment status. For comparison participants, the assessment additionally captured whereabouts and party attendance on October 7. The broader protocol database included 1,268 eligible Nova survivors, of whom 1,167 completed the TP1 online assessment; the remaining 101 did not complete the initial study-entry assessment or did not provide sufficient data for inclusion in that stage. Hence, a total of 1,167 survivors and 508 comparison participants completed this study stage.
- <u>Online probabilistic reinforcement-learning task:</u> The online behavioral task was implemented as a probabilistic two-armed bandit task framed as a fishing game (Aberg et al., 2023). Participants completed three blocks of 20 trials on their smartphones. On each trial, they chose between two lakes, one associated with a 75% reward probability (i.e., successful fishing) and the other with a 25% reward probability. The left-right positions of the lakes were randomized across trials, whereas reward probabilities remained stable within each block. Lake identities were replaced between blocks, requiring participants to relearn reward contingencies for each new stimulus pair.
- <u>Wearable sensors:</u> During TP1, 211 survivors and 113 matched comparison participants attended in-person visits and were recruited for one month of continuous daily-life monitoring using Fitbit Charge 5 devices (Magal et al., 2026). Before and after the monitoring month, participants completed an additional battery of self-report questionnaires and performed a 3-minute electrodermal activity (EDA) measurement session using the Fitbit device.
- <u>Ecological assessments:</u> The one-month monitoring period was accompanied by ecological momentary assessments (EMA) surveys administered every five days, each referring to the preceding five-day period. Surveys assessed substance use, subjective sleep quality, and psychological distress.
- <u>Saliva sampling:</u> Saliva samples were collected during in-person wearable-sensor visits. Two samples were obtained from each participant approximately one hour after their arrival. To reduce potential confounds, participants were instructed to refrain from eating, drinking anything other than water, smoking, or engaging in physical activity for at least one hour prior to their arrival. All samples were collected between 14:00-19:00 and then stored at -20°C and shipped to a specialized saliva-analysis laboratory.
- <u>fMRI session:</u> A subset of 136 participants (73 trauma-exposed participants and 63 comparison participants) underwent a single fMRI session at two acquisition sites: Rambam Health Care Campus (n = 105) and the Weizmann Institute of Science (n = 31). MRI data collection took place between January and September 2024.

#### TP2

- The second assessment time point (June-September 2024) comprised online follow-up questionnaires assessing updated sociodemographic information, employment and economic status, party attendance since the attack, additional crises since the attack, current clinical status, mental-health service utilization, physical health and medications, substance use, sleep quality, attachment, family status before and after the traumatic event, distress and PTSD symptoms. A total of 509 survivors and 99 comparison participants completed this study stage.

#### TP3

- The third assessment time point (March-September 2025 for survivors; August 2025-January 2026 for comparison participants) comprised online questionnaires assessing updated demographics, family status, employment and economic status over the past six months; current clinical status, substance use in the past month, party attendance and significant events in the past six months, mental-health service utilization, and mental state in the context of ongoing regional conflict. TP3 also repeated several TP1 components, including wearable-sensor monitoring, the online probabilistic reinforcement-learning task, ecological assessments, and saliva sampling as described above. Participants additionally completed semi-structured qualitative interviews, a new interoception task and a brief questionnaire regarding location and degree of shelter-seeking during the April 2024 Iranian missile attacks on Israel. A total of 428 survivors and 185 comparison participants completed this study stage.
- <u>Interoception task:</u> The interoception session was completed by 86 Nova survivors and 78 comparison participants. These in-person sessions comprised psychological, cognitive, and physiological blocks, including an interoceptive questionnaire battery, the Heart Rate Discrimination Task (HRDT)(Legrand et al., 2022), and ECG recording during rest, emotionally provoking video viewing, and recovery.
- <u>Semi-structured interviews:</u> Semi-structured qualitative interviews were conducted via Zoom or face-to-face, lasted approximately one hour, and were audio- and video-recorded with consent before transcription. The interview was organized around three chronological segments: neutral introductory conversation and demographics, narrative description of the October 7 event, and post-event recovery. Topics included substance use during the event, time perception, sense of reality, mystical experiences, loss of close others, connections to hostages, duration of trauma exposure, coping since the event, and perceived effects of hostage returns on safety and recovery. Eighty-five (85) survivors completed this study stage between April and November 2025.

### 2.7 Measures

#### Clinical and self-report measures

The primary longitudinal outcomes are PTSD symptom severity, measured with the PTSD Checklist for DSM-5 (PCL-5)(Bovin et al., 2016) and general psychological distress, measured with the Kessler Psychological Distress Scale (K6)(Kessler et al., 2003). PCL-5 items are rated on a 0-4 scale across the DSM-5 PTSD symptom clusters, with a commonly used clinical cut-off of a total score of 33 or above (Blevins et al., 2015; Bovin et al., 2016; Weathers et al., 2013). The K6 includes six items rated on a 0-4 scale, and clinically significant distress was defined as a total score of 13 or above (Furukawa et al., 2003; Kessler et al., 2002; Kessler et al., 2003). For improved transparency, the self-report battery is reported by time point and study stage, as summarized in **Table 1**.

At study entry, participants reported demographics, employment status, pre-event clinical history, prior trauma exposure, mental-health treatment, medications, substance-use history before October 7, detailed substance use during October 7, trauma-exposure severity, peritraumatic control and isolation, social support, social interactions, guilt, sleep quality, PCL-5, and K6. Across subsequent stages, additional measures included the following established self-report inventories: Depression Anxiety and Stress Scale (DASS-21; TP1,2&3)(Lovibond, 1995); Patient Health Questionnaire (PHQ-9; TP1,2&3)(Kroenke et al., 2001); Perceived Stress Scale (PSS; TP1&2)(Cohen et al., 1983); Pittsburgh Sleep Quality Index (PSQI; TP1,2&3)(Buysse et al., 1989); Peritraumatic Dissociative Experiences Questionnaire (PDEQ; TP1&2)(Birmes et al., 2005); Multidimensional Scale of Perceived Social Support (MSPSS; TP1&2)(Zimet et al., 1990); Post-Traumatic Growth Inventory (PTGI; TP1,2&3)(Tedeschi & Calhoun, 1996); Meaning in Life Questionnaire (MLQ; TP1)(Steger et al., 2006); Sense of Coherence (SOC; TP1)(Antonovsky, 1993); Ruminative Response Scale (RRS; TP1)(Treynor et al., 2003); Pain Catastrophizing Scale (PCS; TP1)(Sullivan et al., 1995); Adverse Childhood Experiences scale (ACE; TP1)(Felitti et al., 2019); State Adult Attachment Measure (SAAM; TP1&2)(Gillath et al., 2009); Multidimensional Assessment of Interoceptive Awareness Version 2 (MAIA-2; TP1&2)(Mehling et al., 2018); and a modified version of the Mystical Experiences Questionnaire (MEQ-30, modified version; TP1)(Barrett et al., 2015), adapted to assess substance-related mystical-type experiences before and after the traumatic event. At the interoception session at TP3, participants also completed newly developed study-specific instruments: the Interoceptive Sensibility for PTSD Questionnaire (ISPQ), the Interoceptive Skills Questionnaire (ISQ), and the Body-Substances Questionnaire (BSQ).

Substance-use measures covered three timeframes: (1) use during the October 7 events, including type, dose/amount, timing, perceived strength before and after attack onset, overall helpfulness or disruptiveness; (2) lifetime use before October 7, including typical frequency; and (3) longitudinal post-event use, including type, frequency, purpose of use, perceived helpfulness, co-use patterns, flashback occurrence during use, and change relative to the six months preceding October 7.

#### Wearable sensors

Continuous physiological and behavioral data were collected in daily life during two one-month-long monitoring waves (TP1&3) using Fitbit Charge 5 devices (Fitbit LLC, San Francisco, CA, USA). At TP1, wearable sensor data were available for 211 Nova survivors and 113 comparison participants. A second sampling at TP3 included 104 Nova survivors and 72 comparison participants. Participants wore the device throughout daily activities, including sleep and showering, and received reminders every five days to charge and synchronize the device. If synchronization did not occur, the research team followed up by phone. Sleep data were processed from sleep-onset, offset, and awakening epochs and converted to a 1-minute sleep/awake/unknown vector. Intervals exceeding 30 hours between wake time and the next sleep onset were labelled unknown. Heart-rate samples were recorded every 5-15 seconds and filtered by removing low-confidence samples according to the Fitbit algorithm and values below 40 or above 180 beats per minute, then aggregated to 1-minute resolution. Step count was analyzed at 1-minute resolution; when step data were missing but heart-rate data were present, steps were imputed as zero according to the analytic plan. EDA was recorded using a standardized 3-minute resting protocol after participants washed their hands and sat quietly. Data preprocessing followed established in-house pipelines and wearable-sensor reporting guidelines (Hanuka et al., 2025; Magal et al., 2026; Magal et al., 2022; Simon et al., 2025).

#### Ecological assessments

Parallel to the one-month Fitbit monitoring period, participants completed a short survey every five days. Each survey covered the preceding five-day period and assessed three domains: substance use, subjective sleep quality, and psychological distress. Participants reported the types and quantities of substances used, rated subjective sleep quality on a 0-100 sliding scale, and completed the K6. To maximize compliance, the research team sent a smartphone message every five days instructing participants to synchronize and charge the device and complete the survey. If the device had not been synchronized and/or the survey remained incomplete after 24 hours, a reminder was sent; after 48 hours, a research-team member contacted the participant by phone. This schedule follows ecological-assessment principles while emphasizing the importance of low participant burden.

#### Saliva sampling

Salivary endocrine and inflammatory markers were collected to examine their temporal associations with PTSD symptoms. At TP1, saliva data were available for 185 Nova survivors and 91 comparison participants. A second sampling at TP3 included 106 Nova survivors and 67 comparison participants. Saliva samples were collected using Salivette® cortisol collection swabs (Sarstedt, Nümbrecht, Germany). Swabs were stored at -20 ◦C until analysis. After thawing at 4◦ C, tubes were centrifuged for 10 min at 4000 g to recover the saliva and remove solids. Concentrations of cortisol and alpha-amylase were assayed using a solid-phase enzyme-linked luminescence immunoassay. In addition, samples were assayed using Meso Scale Discovery (MSD) electrochemiluminescence technology for multiplex cytokine detection, focusing on TNF-α, IL-1β, IL-6, and IL-10.

#### Functional MRI

Functional MRI data were acquired from 136 participants at two sites (Rambam Health Care Campus, n = 105; Weizmann Institute of Science, n = 31). Functional scans used a repetition time of 1 second and an isotropic voxel size of 2 x 2 x 2 mm, with continuous respiration and heart-rate recording for physiological monitoring and noise correction. The session comprised: (1) a 6-minute eyes-open baseline resting-state scan; (2) a 6-minute festival-video stress manipulation designed to resemble the Nova festival context without being inherently distressing to the general population; (3) a 6-minute post-stress resting-state scan; (4) a probabilistic reward-learning task lasting approximately 12 minutes following a 12-trial in-scanner training block; and (5) a high-resolution anatomical scan. After the video, participants rated emotional overload, negative affect, trauma-related memory activation, and post-October 7 party attendance. Post-scan tasks included continuous moment-by-moment emotional valence rating of the same video and a knowledge-transfer test assessing retention of reward contingencies learned during scanning.

#### Cardiac interoception

Interoceptive sensitivity and metacognitive awareness were assessed using the Heart Rate Discrimination Task (HRDT)(Legrand et al., 2022), an adaptive Bayesian psychophysical paradigm with interoceptive and auditory exteroceptive control conditions. Participants completed 50 two-interval forced-choice trials in each domain and rated confidence after each trial. During interoceptive trials, heart rate was recorded using a Zephyr Bluetooth-enabled chest-worn device. Trials with reaction times below 100 ms or heart-rate outliers detected using the median absolute deviation rule were excluded. ECG was additionally recorded during three 6-minute phases: resting state, emotionally provoking movie, and recovery. Signals were processed in Python using the Systole package, with peak detection followed by conservative manual correction of missed and ectopic beats.

#### Semi-structured interviews

Interviews were developed specifically to capture the subjective experiences of survivors of the October 7 Nova festival attack. Interviews were conducted via Zoom or face-to-face, lasted approximately one hour, and were audio- and video-recorded with consent before transcription. The interview was structured around three chronological segments: a neutral introductory conversation and demographic information, a narrative account of the event, and the post-event recovery period. Alongside open-ended narrative questions, participants were asked about substance use during the event, time perception, sense of reality, mystical experiences, loss of close others, connections to hostages, duration of exposure, coping since the event, and the perceived effect of hostage returns on safety and recovery. Quantitative ratings included participant-rated emotional valence and arousal at three interview points, interviewer-rated fight/flight/freeze responses, interviewer and transcriber arousal, perceived substance influence and helpfulness, alterations in time perception, positive post-event change, and therapeutic alliance. Prior to their interviews, participants also completed the DASS-21, PHQ-9, K6 and PCL-5 self-report questionnaires.

### 2.8 Data management and quality control

For the PCL-5, exactly one missing item was imputed using the participant median across the remaining 19 items; participants with more than one missing item were excluded from total-score analyses. Uniform responding across all 20 PCL-5 items was flagged as potential non-engagement and excluded from score computation. For K6, total scores were excluded when one or more items were missing. Uniform K6 responding was excluded only when accompanied by uniform responding across the PCL-5 items as well. For visual analogue scales, one missing value was replaced with the scale midpoint, whereas two consecutive missing values or additional missing values were coded as missing according to the prespecified rules. Additional questionnaire-specific scoring and imputation rules were applied for DASS-21, PHQ-9, PDEQ, and MAIA-2, following prespecified code-based rules. Survey metadata, including completion timestamps, response IDs, and completion rates, were retained for duplicate detection and longitudinal data-quality monitoring. All wearable analyses will report on minimum valid-day thresholds, missing-data rules, and modality-specific QC exclusions (Magal et al., 2026). Saliva analyses will report on duplicate handling, outlier exclusion, transformation, assay batch, and plate effects. fMRI analyses will account for acquisition site, head motion, and physiological noise. Interview data were de-identified before transcription/coding, and LLM-based coding will include human validation and privacy safeguards.

### 2.9 Statistical analysis

Primary analyses will examine longitudinal trajectories of PTSD symptom severity (PCL-5) and psychological distress (K6) as a function of time, trauma exposure, and peritraumatic substance-use group. Linear mixed-effects models will include time (modeled as days since October 7), group/substance classification, and their interaction as fixed effects, with participant-level random intercepts and slopes where supported by the data. Substance-use groups will follow the classification used in Netzer et al. (2025), including no use, cannabis/alcohol, MDMA, hallucinogens, stimulants, and polysubstance categories when sample sizes permit (Netzer et al., 2025). Models will adjust for pre-specified covariates such as age, sex, time since event, trauma-exposure severity, prior trauma, treatment exposure, ongoing crises, and relevant modality-specific covariates. Secondary analyses will model PTSD trajectory classes (Simon et al., 2026), wearable-derived sleep and circadian instability (Magal et al., 2026), HR metrics, saliva cytokines and cross-lagged PTSD-inflammation associations, fMRI task/resting-state measures, interoceptive accuracy and metacognition, reinforcement-learning parameters, and interview-derived narrative themes. False-discovery-rate (FDR) correction will be applied within outcome families, and sensitivity analyses will examine attrition, missingness, and robustness of results.

### 2.10 Sample size and power considerations

The study is powered primarily by the natural cohort of Nova festival survivors engaged through SafeHeart and social-media outreach rather than by an a priori randomized design. The broader protocol database included 1,268 eligible Nova survivors, of whom 1,167 completed the TP1 online assessment; 508 comparison participants completed the corresponding TP1 assessment. At TP2, 509 survivors and 99 comparison participants completed the online follow-up, and at TP3, 428 survivors and 185 comparison participants completed the online follow-up. Modality-specific samples included 211 survivors and 113 comparison participants for TP1 and 104 survivors and 72 comparison participants for TP3 wearable monitoring and ecological assessments; saliva data from 185 survivors and 91 comparison participants at TP1 and from 106 survivors and 67 comparison participants at TP3; 136 fMRI participants (73 survivors, 63 comparison participants); 164 interoception participants (86 survivors, 78 comparison participants); and 85 completed semi-structured interviews, of which 83 are currently included in the analytic interview sample. The published PTSD-trajectory analysis included 392 survivors with usable PCL-5 data across two waves (Simon et al., 2026). Final analysis-ready samples will be defined separately for each modality according to modality-specific quality-control and missing-data rules.

## 3. Discussion

The Nova Protocol is among the most comprehensive longitudinal, multimodal investigations of civilian mass-trauma cohorts following a single, time-locked traumatic event. By integrating online self-report questionnaires, prolonged daily-life physiological and behavioral monitoring using wearable sensors, ecological follow-up surveys, saliva-based endocrine and inflammatory markers, structural and functional MRI, interoception, reinforcement learning, and qualitative interviews across a two-year window, the protocol is positioned to examine how symptoms, behavior, physiology, inflammation, neural function, interoceptive processing, and subjective trauma phenomenology jointly shape post-trauma trajectories. The value of this protocol is already reflected in initial Nova cohort outputs, which have characterized peritraumatic psychoactive-substance exposure and early clinical outcomes (Netzer et al., 2025), circadian instability as a prospective marker of PTSD risk (Magal et al., 2026), and adverse PTSD symptom trajectories during the first post-trauma year (Simon et al., 2026). The present protocol paper situates these initial findings within the broader longitudinal and multimodal framework from which they emerged.

Several design features are notable strengths of the Nova protocol. First, the narrow exposure window reduces heterogeneity in trauma timing, a common limitation of PTSD studies. Second, the sociocultural comparison group helps isolate trauma-specific effects from characteristics of trance-community participation and substance-use culture. Third, the wearable and ecological follow-up components capture sleep-wake behavior, HR, activity, substance use, sleep quality, and distress in daily life rather than relying exclusively on retrospective questionnaires (Magal et al., 2026; Magal et al., 2022; Simon et al., 2025). Fourth, saliva sampling enables examination of endocrine and immune/inflammatory mechanisms and their temporal relations with PTSD symptoms. Fifth, qualitative interviews provide phenomenological information that questionnaires cannot capture, including time perception, sense of reality, mystical experiences, social connection and evolving coping. Finally, the multimodal nature of the protocol permits cross-level triangulation across subjective, behavioral, physiological, immune, and neural domains. The protocol also has methodological implications for ecological assessment after trauma. The five-day survey schedule is less intensive than classic high-frequency EMAs, but it may be more feasible in a highly distressed population while still capturing repeated changes in substance use, sleep quality, and distress during real-world recovery.

Several limitations should also be acknowledged explicitly. The cohort is self-selected through support groups, peer referrals, and social media, and may underrepresent survivors with the most severe psychopathology or those who disengaged from support networks. Substance use during the attack was self-reported retrospectively and is vulnerable to recall bias, social-desirability effects, and confounding by pre-existing differences between substance-use groups. Because the study is observational and substance exposure was not randomized, analyses cannot establish causal effects of any of the consumed substances. In addition, attrition across repeated assessment time points and intensive in-person modalities may introduce selection bias, and sample sizes differ across modalities. For example, the fMRI subsample was necessarily more selective because of MRI contraindications. Longitudinal analyses will therefore use analysis-specific samples defined by available repeated data and quality-control criteria. Furthermore, the ongoing war and additional crises after October 7 are both clinically meaningful and analytically challenging, because they may influence symptom trajectories and biological measures. In this respect, although comparison participants were not directly exposed to the Nova attack, they were also embedded in the same broader national context of ongoing war and may have experienced indirect, vicarious, or subsequent stress exposure. Finally, consumer wearables provide scalable ecological data but are less precise than laboratory polysomnography or ECG-grade monitoring, requiring transparent quality-control and validation procedures.

Taken together, the Nova Protocol provides a rare, ethically non-replicable naturalistic framework for studying how clinical symptoms, daily-life physiology, inflammation, neural function, interoception, reinforcement learning, and subjective trauma phenomenology jointly shape response trajectories after civilian mass trauma. Findings should inform early risk detection, individualized monitoring, and post-disaster mental-health care, while also informing models of peritraumatic pharmacology and mechanisms relevant to MDMA- and psychedelic-assisted interventions for PTSD.

## Ethics approval and consent to participate

The study was approved by the University of Haifa Institutional Ethics Committee (approval no. 374/23) and the Rambam Health Care Campus Institutional Review Board (RMB0553-23). All participants provided informed consent prior to participation at each assessment time point.

## Data availability statement

Because the dataset contains highly sensitive trauma, health, substance-use, biological, neuroimaging, and interview data, de-identified individual-level data cannot be made publicly available without additional ethical approval and data-use agreements. Requests for access to de-identified data or analysis code will be considered by the corresponding author, subject to institutional approval and participant privacy constraints.

## Author contributions

R.A: conceptualization, methodology, data collection, writing-original draft, supervision, funding acquisition.

O.N: methodology, data collection, formal analysis, data curation, writing-review and editing, project administration.

N.M: methodology, data collection, formal analysis, data curation, project administration.

L.S: methodology, data collection, formal analysis, data curation, writing-review and editing, project administration.

A.H: formal analysis, data curation, project administration.

M.O: methodology, data collection, data curation.

O.R: methodology, data collection, formal analysis, data curation.

S.C: methodology, data collection, project administration.

M.B: data collection, data curation.

M.G: data collection, data curation, project administration.

R.M: data collection, formal analysis, data curation.

Y.S: methodology. N.M: data collection.

A.S: methodology, data collection, formal analysis, data curation.

E.E: data collection, data curation.

D.S: methodology, data collection, formal analysis, data curation.

T.P: formal analysis, data curation.

R.G: conceptualization, writing-review and editing.

R.S: conceptualization, methodology, data collection, writing-review and editing, supervision, project administration, funding acquisition.

## Acknowledgements

We are deeply grateful to the Nova survivors and participants for their time, trust, and willingness to contribute to this research during an exceptionally challenging period in their lives. Their courage and generosity in sharing their experiences make this work possible and represent an invaluable contribution to advancing our understanding of trauma and recovery. We also thank SafeHeart for facilitating survivor outreach and support, including the SafeHeart NGO management and early volunteers Reut Plonsker, Karina Dessau, Tal Zagursky, Yair Grynbaum, Nir Tadmor, Guy Simon, Igal Tartakovsky, Irit Hacmun, Shiran Maor, Dr. Demian Halperin, and Efrat Atun, who worked tirelessly to ensure support and treatment for survivors. We thank all SafeHeart volunteers, clinicians, and members of our research team. The long author list reflects the collaborative nature of this protocol and the collective effort required to design and execute it. Additional team members that should be acknowledged include Atalya Ceder, Shai Sayegh, Nitsan Sagi, Netta Druckman, Shira Asher, Shani Berkovitch, Gal Markovitch, Shiraz Azulay, Yonatan Zairi, Kristoffer Aberg, Tomer Cohen, Bar Kroiter, Nir Aviram, and Shaked Tzivin. Finally, we thank colleagues and collaborators for their continuous support and valuable consultation, including Dr. Pia Rothstein, Prof. Rony Paz, Prof. Ayelet Eran, Dr. Edna Furman-Haran, Prof. Nicolas Rohleder, Dr. Dimitris Repantis and Prof. Uri Hertz.

